# Diagnostic accuracy of a novel SARS-CoV-2 antigen-detecting rapid diagnostic test from standardized self-collected anterior nasal swabs

**DOI:** 10.1101/2021.04.20.21255797

**Authors:** Bilgin Osmanodja, Klemens Budde, Daniel Zickler, Marcel G. Naik, Jörg Hofmann, Maximilian Gertler, Claudia Hülso, Heike Rössig, Philipp Horn, Joachim Seybold, Stephanie Lunow, Melanie Bothmann, Astrid Barrera-Pesek, Manuel Mayrdorfer

## Abstract

**Background:** Antigen-detecting rapid diagnostic tests (Ag-RDT) for SARS-CoV-2 offer new opportunities for the quick and laboratory-independent identification of infected individuals for control of the SARS-CoV-2 pandemic. Despite the potential benefits, nasopharyngeal sample collection is frequently perceived as uncomfortable by patients and requires trained healthcare personnel with protective equipment. Therefore, anterior nasal self-sampling is increasingly recognized as a valuable alternative.

**Methods:** We performed a prospective, single-center, point of care validation of an Ag-RDT using a polypropylene absorbent collector for standardized self-collected anterior nasal swabs. Real-Time Polymerase Chain Reaction (RT-PCR) from combined oropharyngeal/nasopharyngeal swabs served as a comparator. Primary endpoint was sensitivity of the standardized Ag-RDT in symptomatic patients with medium or high viral concentration (≥ 1 million RNA copies on RT-PCR for SARS-CoV-2).

**Results:** Between February 12 and March 22, 2021, 388 participants were enrolled. After exclusion of 9 patients for which no PCR result could be obtained, the novel Ag-RDT was evaluated based on 379 participants, of which 273 were symptomatic and 106 asymptomatic. In 61 samples from symptomatic patients with medium or high viral load (≥ 1 million RNA copies), the sensitivity of the standardized Ag-RDT was 96.7% (59/61; 95%CI: 88.7-99.6%) for the primary endpoint. In total, 62 positive Ag-RDT results were detected out of 70 RT-PCR positive individuals, yielding an overall sensitivity of 88.6% (95%CI: 78.7-94.9%). Specificity was 99.7% (95%CI: 98.2-100%) in 309 RT-PCR negative individuals.

**Conclusion:** Here, we present a validation of a novel Ag-RDT with a standardized sampling process for anterior nasal self-collection, which meets WHO criteria of ≥80% sensitivity and ≥97% specificity. Although less sensitive than RT-PCR, this assay could be beneficial due to its rapid results, ease of use, and suitability for standardized self-testing.

(Funded by Drägerwerk AG & Co. KGaA, Lübeck, Germany; ClinicalTrials.gov number NCT04698993)

## Background

Various antigen-detecting rapid diagnostic tests (Ag-RDTs) for SARS-CoV-2 are now commercially available.^1^ Performed in an appropriate indication, they can support rapid decisions with respect to isolation, contact tracing, and treatment of patients with Covid-19.^2^ Since nasopharyngeal (NP) swabs are frequently perceived as uncomfortable by patients and must be collected by trained healthcare personnel, they are of limited use when establishing a population wide testing strategy.^3^ Fortunately, there is an increasing evidence base supporting the use of alternative sampling methods, including anterior nasal self-collection.^4, 5^ This easier sample collection method can aid to achieve higher reliability of Ag-RDTs for self-testing.

The primary objective of this prospective diagnostic accuracy study was to assess sensitivity and specificity for a novel Ag-RDT with supervised, self-collected anterior nasal swab sample using a porous polypropylene absorbent collector against the reference standard RT-PCR collected from an oropharyngeal (OP)/NP swab.

## Methods

The study protocol was approved by the ethical review committee of the federal state of Berlin and registered under ClinicalTrials.gov (NCT04698993). All experiments on human subjects were performed in accordance with the Declaration of Helsinki, implying that all participants provided informed consent. The study took place at two ambulatory SARS-CoV-2 testing facilities at Charité - Universitätsmedizin Berlin, Germany, from February 12 to March 22, 2021. At study site A, symptomatic adults suspected of SARS-CoV-2 infection were enrolled, while at study site B, asymptomatic and symptomatic employees and students were enrolled, participating in the regular hospital surveillance scheme. Main inclusion criterion for symptomatic patients was onset of COVID-19 symptoms within 7 days prior to testing. Main exclusion criteria were bleeding disorder, nasal spray application before testing, pregnancy and breastfeeding. Complete inclusion and exclusion criteria are provided in Table 1. Only participants with both an evaluable test result for the Ag-RDT and the RT-PCR reference standard were included in the analysis.

**Table 1:** Inclusion and exclusion criteria.

|  |  | Inclusion Criteria | Exclusion Criteria |
| --- | --- | --- | --- |
| General |  | • $\geq 18$ years old<br>• Written informed consent<br>• Pre-existing need for SARS-CoV-2 testing, i.e.<br>○ COVID-19 symptoms<br>○ Known or suspected exposure to SARS-CoV-2<br>○ Screening | • $< 18$ years old<br>• Unable to provide informed consent<br>• Pregnant or breast-feeding women<br>• Involuntarily held in an institution<br>• Bleeding disorder<br>• Application of nasal spray prior to testing on the day of testing<br>• Hospitalization/ inpatient treatment |
| Specific | Asymptomatic | • Asymptomatic during the previous 14 days | • COVID-19 symptoms during the previous 14 days |
|  | Symptomatic | • COVID-19 symptom(s) present on the day of testing<br>• Symptoms started 0-7 days prior to testing day | • Symptoms started more than 7 days prior to testing day |

First, participants received a combined OP/NP swab (eSwab from Copan with 1mL Amies medium) as per institutional recommendations for RT-PCR. Subsequently, participants underwent an instructed, self-collected bilateral anterior nasal swab for the Ag-RDT (Dräger Antigen Test SARS-CoV-2 by Dräger Safety AG & Co. KGaA, Lübeck, Germany). The test comes as a one-piece test kit comprising a test cassette and a removable sample collector. The tip of the sample collector is a rigid, porous polypropylene sponge, which is used to swab the anterior nose and collect mucus and epithelial cells. After the general test procedure was explained to participants, verbal instruction was given to blow the nose once with a tissue. Next, the participants inserted the absorbent collector vertically 2–3 cm into the nostril and wiped the nasal walls in a circular motion for 30 seconds (Figure 1). This sampling process was repeated in the other nostril. After sampling, the sample collector was inserted and locked in the test cassette, where samples were analyzed immediately (within 15 min) after sampling at point-of-care by study physicians according to the manufacturer’s instructions. The test uses the lateral flow assay principle detecting SARS-CoV-2 nucleocapsid protein with visual read-out after 15–20 minutes. The test cassette is a self-contained unit, which allows sample analysis while avoiding further contact with potentially infectious material and making handling of additional liquids, e.g. pipetting or dropping buffers, unnecessary.

**Figure 1:**
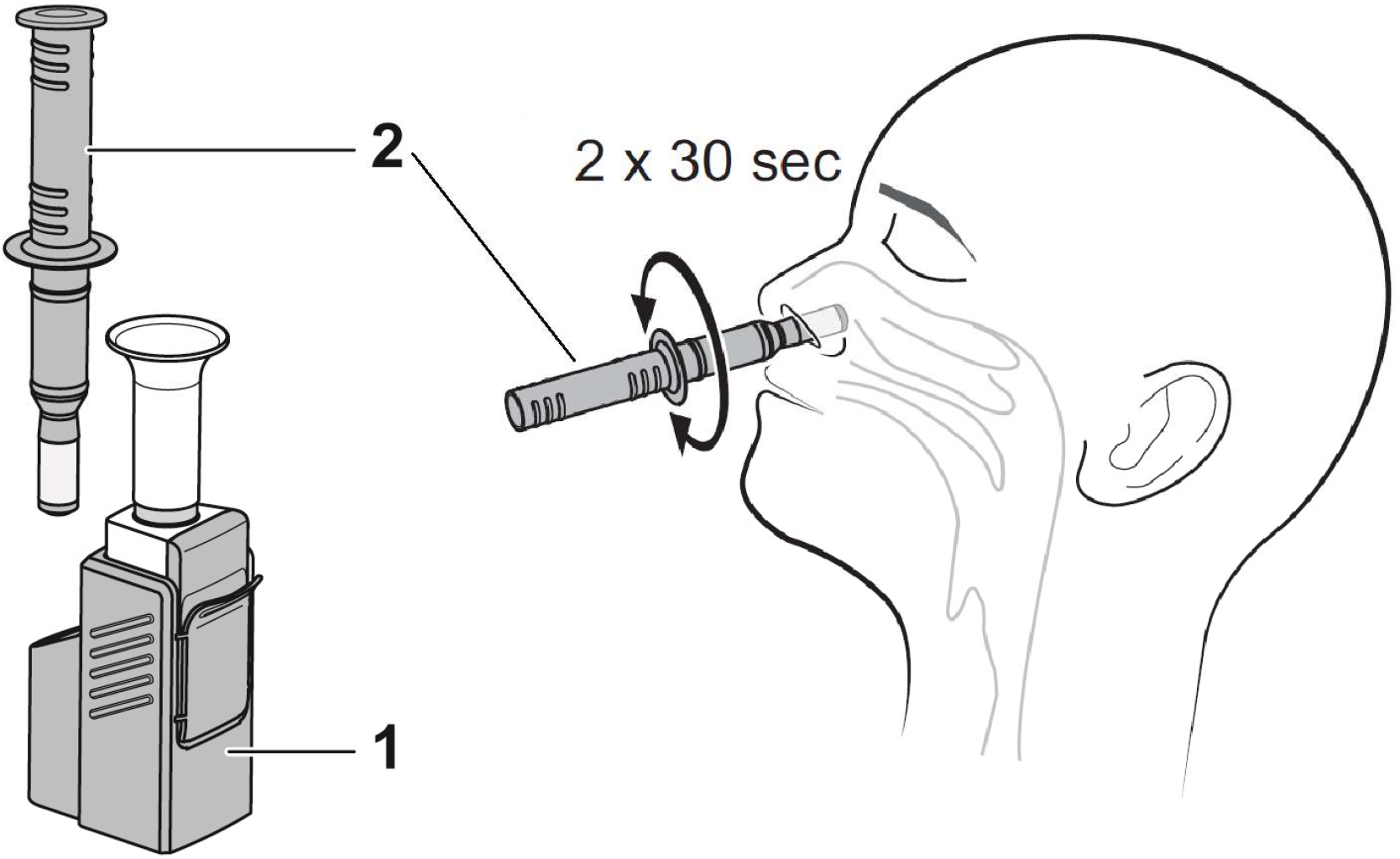
Sample collection - on the left, the test kit comprising a test cassette (1) and a removable sample collector (2) are shown. On the right, the sampling procedure is shown schematically.

The Roche Cobas SARS-CoV-2 assay (Pleasanton, CA, United States) was performed on a cobas® 6800/8800 analyzer (Roche Diagnostics, Mannheim, Germany) targeting both, *orf1a/b* (SARS-CoV-2) and *E*-gene (pan-Sarbecovirus). Viral concentration was classified into 3 categories (low: < 1 million, medium: 1 – 10 million, high: > 10 million RNA copies) based on standard preparations provided by Institute of Standardization, Düsseldorf, Germany. For all first-time diagnosed SARS-CoV-2 samples, typing for variants of concern (VoC) was performed, according to Phylogenetic Assignment of Named Global Outbreak (PANGO) Lineages classification.^6^

Primary endpoint was sensitivity of the novel Ag-RDT in symptomatic patients with medium or high viral concentration (≥ 1 million RNA copies) on RT-PCR.^7-9^

Secondary endpoints were overall sensitivity and specificity, specificity in asymptomatic patients and frequency of nosebleed or unbearable pain due to specimen collection.

Data analysis was performed using the statistical programming language R, version 3.6.3.^10^ The R software package ggplot2 was used for data visualization.^11^ Sensitivity and specificity were determined, and the corresponding confidence intervals were calculated using the Clopper-Pearson method. Two-proportions Z test was used for group comparison based on mutation status.

The study was stopped early, due to poor recruitment and Sponsor decision after 70 RT-PCR positive participants.

## Results

Of 422 patients invited, 388 (91.9%) consented to participate. Nine Ag-RDT negative participants (n=7 asymptomatic, n= 2 symptomatic) were excluded as no RT-PCR result could be obtained until the end of the study due to data protection issues. For another symptomatic patient, RT-PCR found RNA at the limit of detection, but repeated RT-PCR testing was negative, hence the patient was classified as RT-PCR negative. In summary, 379 patients were included into the final analysis, of which 273 were symptomatic and 106 were asymptomatic (Figure 2).

**Figure 2:**
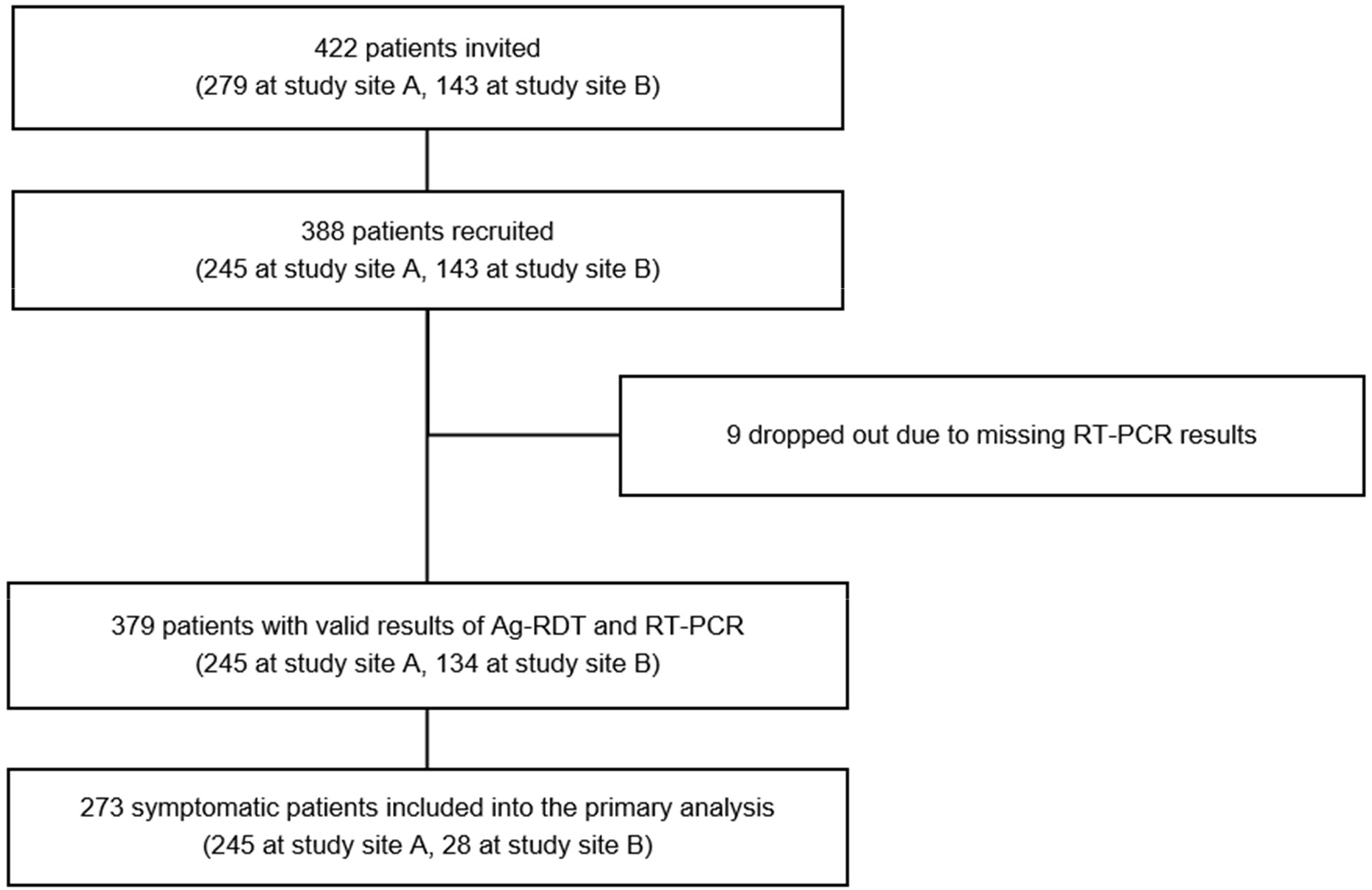
Participant Flow Diagram

The average age of participants was 34.0±10.8 years with 53.3% female and 46.7% male. Among all participants, 14.0% had comorbidities. Duration of symptoms at the time of presentation was on average 2.8±1.8 days among the 273 symptomatic patients. Among all 379 participants, 70 (18.5%) tested positive for SARS-CoV-2 RNA, one of which was asymptomatic.

### Primary endpoint

In 61 symptomatic participants with medium or high viral concentration (≥1 million RNA copies), the sensitivity of the Ag-RDT was 96.7% (59/61 RT-PCR positives detected; 95% CI 88.7% - 99.6%). In 9 patients with low viral concentration (<1 million copies) only 3 tested positive with the Ag-RDT test (Table 2).

**Table 2:**
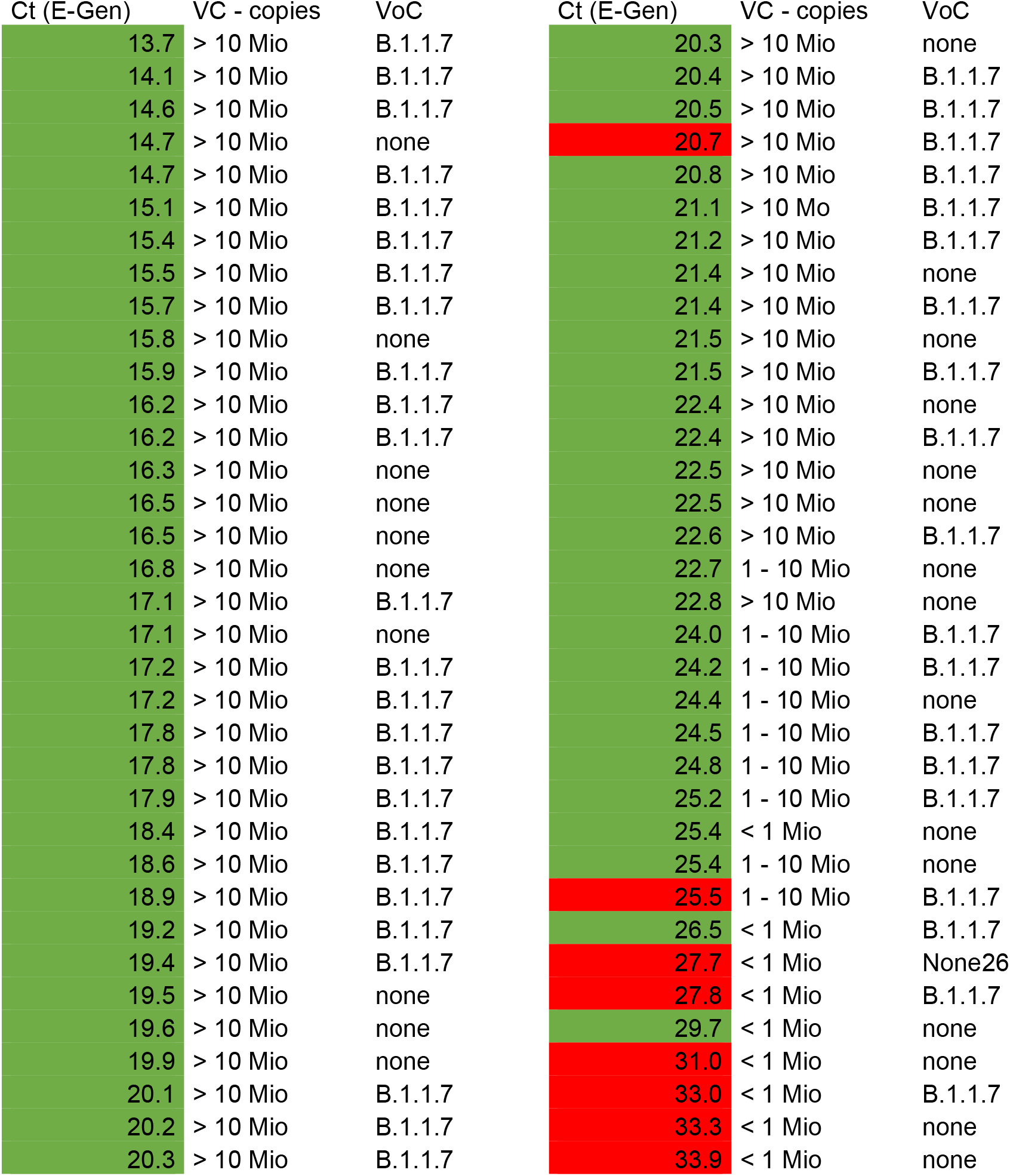
Antigen-detecting RDT results with a supervised self-collected anterior nasal swab in 70 RNA positive patients from combined oropharyngeal/nasopharyngeal swab. Abbreviations: Ct – cycle threshold, VC – Viral concentration, VoC – Variant of Concern. Green – positive Antigen-detecting rapid diagnostic test. Red – negative Antigen-detecting rapid diagnostic test.

### Secondary endpoints

In total, the novel Ag-RDT showed a sensitivity of 88.6% (62/70 PCR positives detected; 95% CI 78.7% - 94.9%) as shown in Figure 3. Overall, specificity was 99.7% (308/309 RNA negatives detected; 95% CI: 98.2% - 100%) compared to RT-PCR.

**Figure 3:**
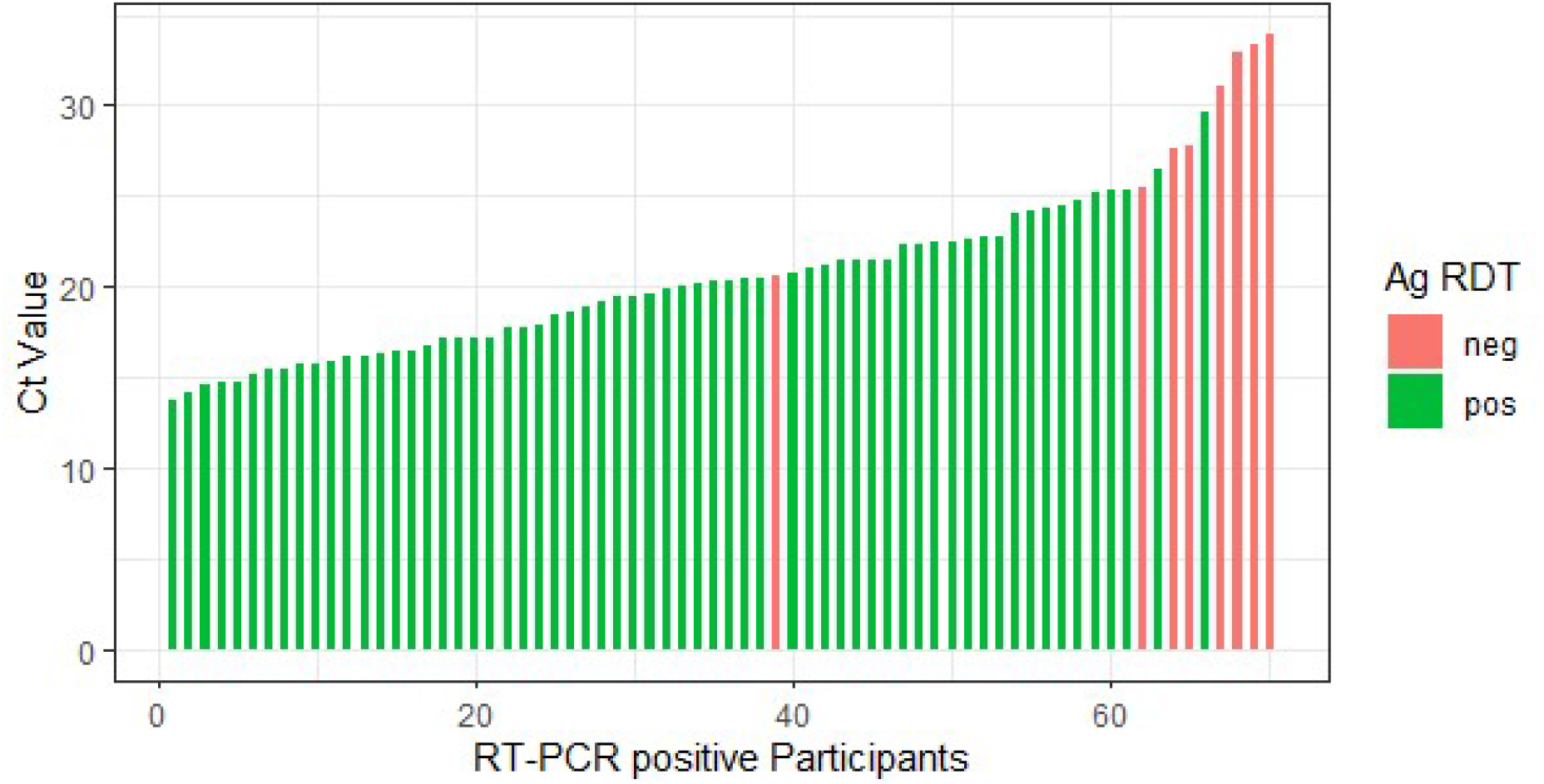
Bar plot showing Ag-RDT results and the corresponding Ct-values of 70 RT-PCR positive patients. Ct – Cycle threshold, Ag RDT (Antigen rapid diagnostic tests), neg – negative, pos – positive, RT-PCR (Real-Time Polymerase Chain Reaction).

Among the 106 asymptomatic participants, one tested positive on RT-PCR with a Ct-value of 31, but was negative on Ag-RDT. For the remaining 105 asymptomatic participants, who tested negative on RT-PCR, specificity of the Ag-RDT was 100%.

Regarding mutation status, 44 participants were diagnosed with VoC B.1.1.7 and for the remaining 26 participants no VoC was found. Sensitivity of Ag-RDT among those patients with VoC B.1.1.7 was 88.6% (39/44) and did not differ significantly (p=0.9075) from those without VoC, where sensitivity was 84.6% (22/26).^6^

### Safety and usability

During the study, no adverse events and no invalid results on Ag-RDT occurred. Regarding usability, comfort and safety, 4.2% (16/379 participants) reported light pain during the self-collection, but none reported strong or unbearable pain, and none developed nosebleed.

## Conclusion

In this study, we demonstrated diagnostic accuracy of a novel Ag-RDT from self-collected anterior nasal swab, meeting the WHO criteria of ≥80% sensitivity and ≥97% specificity.^2^ Hereby, we follow other authors, who have demonstrated that supervised self-sampling from the anterior nose is a reliable option for Ag-RDT, yielding diagnostic accuracies comparable to those from nasopharyngeal swabs.^4^

In comparison to other Ag-RDTs, the standardized absorbent collector used in this study is a rigid, porous sponge, which might reduce variability in sampling method. While Ag-RDTs are rather reliable, the Achilles heel of testing, and in particular of anterior nasal swabs, is sampling procedure. Most Ag-RDTs for self-testing rely on flexible specimen collectors that can be used to perform anterior nasal, mid-turbinate or nasopharyngeal sample collection. In contrast, using a standardized sampling procedure designed for anterior nasal testing may result in less variability, which is of utmost importance for the reliability of self-testing. Additionally, since the test comes as self-contained device comprising all required biochemical reagents and the dilution buffer to carry out the test, the risk of contamination in the case of supervised self-testing is minimized for the testing personnel.

When compared to results from other test accuracy studies, the overall sensitivity of 88.6% found in our study is higher than the one reported in a recent systematic review, where the average sensitivity of Ag-RDTs in the first week after symptom onset was 78.3%.^12^ Among those patients with medium or high viral concentration, sensitivity in our study was 96.7%, which is comparable to the average sensitivity of 94.5% among patients with Ct values of ≤25 reported in the same review.^12^ Among 106 asymptomatic participants, which we included in our study, only one tested positive on RT-PCR with a Ct-value of 31, but was negative on Ag-RDT. The remaining 105 participants were correctly tested negative, which together with the overall sensitivity of 99.7%, makes positive results of this Ag-RDT highly reliable. Still, this emphasizes the importance of patient selection for Ag-RDT. While Ag-RDT are of high sensitivity in the first week of disease, their sensitivity in the second week or among asymptomatic patients is only 51.0% or 58.1%, respectively.^12^

Limitations of our study arise due to the fact, that OP/NP testing for RT-PCR was performed before Ag-RDT due to organizational reasons at the testing facility. Theoretically, this could transfer virus from the nasopharyngeal space to the anterior nose, but seems of little practical relevance since all patients were instructed to blow their nose to increase viral load in the anterior nose in any case. Another limitation in our and other studies is that Ct-values and viral concentration estimation are highly dependent on sample quality. In our study, experienced medical staff performed sampling and further processing was almost identical for all samples. We used a cutoff of 1 million copies for the primary endpoint for two reasons. First, an internal standard of 1 million RNA copies was tested as a one-point calibration with each PCR run. Second, in addition to technical reasons, viral concentrations below 1 million RNA copies indicate a lack of contagiosity in the late phase of Covid-19, and can be used to guide isolation measures according to German healthcare authorities.^7-9^

Considering the ease-of-use of Ag-RDTs, self-sampling and patient self-testing is the main future use case for such tests. The standardized sampling and test procedure of the Ag-RDT investigated in this study may allow for more reliable self-testing, which can increase testing frequency and can have significant impact on the pandemic.

## Data Availability

Available data: (1) De-identified data that underlie the results in this paper. (2) Analysis code. Until 5 years after publication. Available to: researchers who provide a sound proposal and all study sites and the Sponsor agree to sharing the data. Proposals should be directed towards the corresponding author.

## Acknowledgements

We thank Sabrina Pein, Stefan Kruska, Silke Kasbohm, Nicole Walendowski and Rene Nadolny for their support.

Marcel G. Naik is participant in the BIH-Charité Digital Clinician Scientist Program funded by the Charité – Universitätsmedizin Berlin and the Berlin Institute of Health.

## Author contributions

BO and MM designed the study and organized the study sites. KB and MG contributed to the study design. BO, KB, MGN, and DZ enrolled patients, and performed the investigation. HR, MG, CH, and JS coordinated and supervised the testing facilities. SL, MB, PH, ABP ran the testing facilities and performed the comparative investigation. BO and MM led the data analysis. JH was responsible for PCR testing and contributed to the interpretation of the data. DZ was the principle investigator of the study. BO, KB, MM, and JH wrote the manuscript. All authors have reviewed the final manuscript.

